# Structural Variant Imputation in Samoans Using a Population-Specific Reference Panel

**DOI:** 10.64898/2026.07.07.26357461

**Authors:** Lauren M. Spor, Emily M. Liau, Alba Sanchis-Juan, Andrew N. Silva, Hong Cheng, Take Naseri, Muagututi‘a Sefuiva Reupena, Satupa‘itea Viali, John Tuitele, Erin E. Kershaw, Ranjan Deka, Nicola L. Hawley, Stephen T. McGarvey, Daniel E. Weeks, Jenna C. Carlson, Harrison Brand, Ryan L. Minster

**Author notes:** These authors contributed equally to this work (Jenna C. Carlson, Harrison Brand, Ryan L. Minster).

## Abstract

Structural variants (SVs) are often excluded from genetic research because they are difficult to call, but they can have substantial effects on phenotypic traits. SVs have not previously been characterized in Samoans, an understudied population with a high burden of complex diseases. Using short-read whole genome sequencing data, we called SVs in 1,276 Samoans and created a Samoan-specific imputation panel inclusive of both SVs and single nucleotide variants (SNVs), called the *Soifua Manuia-SV* panel. Using this panel, we imputed SVs and SNVs in 3,611 Samoans with array data, enabling analysis of SV-phenotype associations in a sample of 4,887 Samoan participants. We evaluated imputation performance in Samoans against two other reference panels: (i) an SNV-only Samoan-specific reference panel, to assess whether SV inclusion impacts SNV imputation, and (ii) an SV and SNV, multi-ancestry reference panel composed of 1000 Genomes participants, which did not include Polynesians, to assess the importance of including the target population in the reference panel. The *Soifua Manuia-SV* panel substantially outperformed the multi-ancestry SV and SNV panel, yielding 5.5 million more high-quality (r^2^ ≥ 0.8) variants, including over 8,000 more high-quality SVs. SNV imputation based on the two Samoan-specific panels performed similarly overall, suggesting that SV inclusion does not strongly impact SNV imputation quality. This work highlights the importance of population representation for accurate imputation.

## Introduction

Structural variants (e.g., deletions, duplications, insertions, inversions, and translocations) are a major source of human genetic variation but are often excluded from most genetic research due to their size (at least 50 base pairs long), which makes them difficult to call. However, structural variants (SVs) can play a significant role in both complex and monogenic disorders through mechanisms such as altering gene dosage, creating gene fusions, and disrupting reading frames, warranting their inclusion in genomic research.^1,2^

Researchers can leverage existing short-read, whole genome sequencing (WGS) to detect SVs using pipelines such as GATK-SV, which can capture approximately 7,000 to 10,000 SVs per genome.^3,4^ Long-read sequencing can detect more than twice as many SVs per genome than short-read sequencing, but it is much more costly.^5–7^ SV imputation from a publicly available reference panel offers a resource-efficient alternative: rather than generating new data, it compares existing participant genotypes to reference haplotypes and uses the closest matches to infer missing genotypes statistically.

There are few genome-wide SV imputation panels available online. Noyvert et al. created a publicly-available panel using long-read sequencing from 888 participants sampled from the 1000 Genomes Project (1000G), which contains individuals of admixed American, African, East Asian, European, and South Asian ancestries. To assess imputation accuracy, Noyvert et al. performed leave-one-out imputation for each of the 888 participants. After demonstrating sufficient SV imputation quality, they successfully imputed SVs into over 488,000 UK BioBank participants, a sample which is largely White European.^7^ Similarly, Fan et al. created a SV imputation panel containing long-read sequencing data from 943 Han Chinese individuals, which is hosted on an online server but is not available for download. They demonstrated that genetic similarity is crucial for good imputation quality, as their panel imputed 4.3-fold more high-quality SVs in Han Chinese individuals compared to a 3,202-person 1000G panel.^8^

There have yet to be studies examining SVs in Samoans, a Polynesian group that is underrepresented in biomedical research and who face a high burden of complex diseases such as obesity, diabetes, and cardiovascular disease. This study describes a resource that can bolster representation of Polynesian populations in genetic research, thereby enhancing overall diversity and enabling opportunities to study population-specific SVs that may be important in understanding genetic drivers of complex diseases.

We previously created a Samoan-specific reference panel containing only single nucleotide variants (SNVs) and demonstrated that Samoan imputation is most accurate when Samoans are represented in the reference panel.^9^ Here, we identify SVs using existing short-read WGS data in 1,276 Samoans, create an SV-inclusive Samoan-specific reference panel, and impute SVs and SNVs into over 3,600 non-sequenced Samoans. To determine the extent to which SV inclusion and Samoan representation impacts imputation quality (r^2^), we compared imputation metrics between this new panel, its SNV-only predecessor, and the multi-ancestry 1000 Genomes SV panel.^7,9^ We found that, much like the previously developed SNV panel, a reference panel that includes SVs measured in Samoan participants substantially outperformed a reference panel composed of genetically dissimilar individuals.

## Methods

### Participants

For studies related to cardiometabolic health, we previously recruited 4,887 unique adult participants across the U.S. Territory of American Samoa and the Independent State of Samoa (Samoa) during the following years: 1990, 1991, 2002, 2003, and 2010 (Figure 1).^10–14^ Of the participants, 17% were recruited from American Samoa, and 58% of the participants were female (*n* = 2,825).

**Figure 1:**
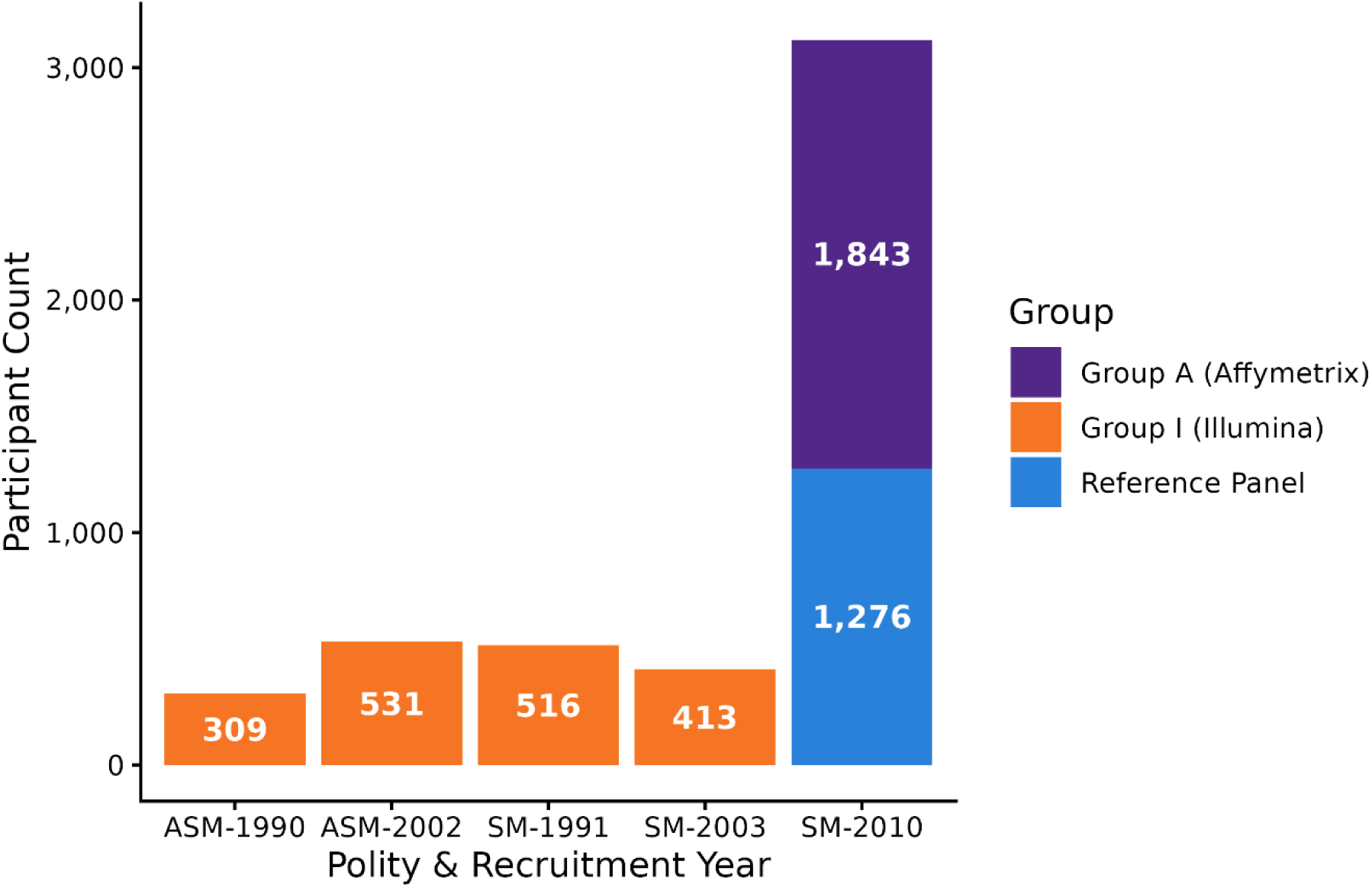
Participants grouped by polity and recruitment year. Samoan participant counts grouped by polity and recruitment year and colored by imputation group/array or reference panel. Key: ASM = American Samoa; SM = Samoa. Note: There is one cross-cohort duplicate between Group A and Group I which was maintained for concordance metrics. There are 4,887 unique participants across the reference panel, Group A, and Group I. See Methods: Imputation Concordance for detailed information.

In 1990 and 1991, we recruited 825 participants (ages 25 to 55 years old) from Samoa and American Samoa into a longitudinal study of adiposity and cardiovascular risk. Of these participants, 309 were recruited from American Samoa in 1990, and 516 were recruited from Samoa in 1991. All participants denied a history of heart disease, hypertension, or diabetes at the time of recruitment, and they reported having four Samoan grandparents as part of study eligibility, an empirically validated reflection of their ancestry.^10,11,15^

In 2002 and 2003, we recruited 944 participants from Samoa and American Samoa as part of a family study of cardiometabolic traits. Of these participants, 531 adults (ages 18 to 89 years old) were recruited from American Samoa in 2002, and 413 adults (ages 19 to 82 years old) were recruited from Samoa in 2003. All participants reported Samoan ancestry, but they were not required to have four Samoan grandparents as part of study eligibility.^12,13^

In 2010, we recruited 3,119 participants (ages 24.5 to 65 years old) across 33 villages in Samoa to participate in cardiometabolic and obesity genetic research. One participant was subsequently identified to have also been a participant in the 1990 study in American Samoa. We retained data from both of their samples for concordance metrics. Participants self-reported Samoan ancestry by having four Samoan grandparents. Comprehensive details about sample selection and representativeness are described in Hawley et al., 2014.^14^

### IRB and Consent

Research protocols, informed consents, and secondary analyses of these studies were approved by the Health Research Committee of the Samoan Ministry of Health and the Institutional Review Boards of Brown University, the University of Cincinnati, and the University of Pittsburgh for the 1990, 1991, 2002, 2003, and 2010 cohorts and additionally the American Samoa Department of Health Institutional Review Board for the 1990, 1991, 2002, and 2003 cohorts.^10–14^ All participants were informed about their rights verbally in Samoan by trained research staff before obtaining their written consent.

### Genotyping

We previously genotyped participants using either the Affymetrix 6.0 or Illumina Global Screening arrays, containing 894,008 and 634,756 typed SNVs on chromosomes 1 through 22 and X, respectively. Participants recruited in 1990, 1991, 2002, or 2003 were genotyped using the Illumina Global Screening array (*n* = 1,769), whereas participants recruited in 2010 were genotyped using the Affymetrix array (*n* = 3,119). We previously conducted extensive quality control using a pipeline developed by Laurie et al.; additional details are provided in Minster et _al._16,17

We generated INFOSTIP rankings using the Affymetrix array data to select 1,285 participants who best represented the Samoan cohort, then Trans-Omics for Precision Medicine (TOPMed) performed WGS for these participants.^18,19^ All genotypes are in the human genome build hg38.

### Samoan Genotypes Compared to Other Population Groups

To visualize where Samoans cluster compared to other population groups, we compared the aforementioned Samoan genotypes to 2,504 genotypes from the multi-ancestry 1000G cohort, which were downloaded from TOPMed.^19,20^ The 1000G participants comprise 2,504 individuals labeled admixed American (*n* = 347), African (*n* = 661), East Asian (*n* = 504), European (*n* = 502), and South Asian (*n* = 489), with one additional participant labeled as African and European. Following the TOPMed Analysis Pipeline, we used KING, PC-AiR, and PC-Relate to create kinship-adjusted principal component plots.^21–24^ We also generated admixture plots using ADMIXTURE v1.3.1 to visualize Samoan population structure compared to other population groups.^25^ The admixture analysis used 13,400 SNVs that were genotyped across all Samoan and 1000G participants, were more than 200 kb apart, and had a minor allele frequency (MAF) > 0.01. Cross-validation error was calculated for *K* = 1 to *K =* 6 ancestral populations to identify an optimal number of clusters. The cross-validation error decreased sharply from *K* = 1 to *K* = 4, then declined gradually to *K* = 6, so *K* = 6 was selected to describe the genetic structure across Samoan and 1000G participants.

### Structural Variant Calling

We used the default settings of GATK-SV v0.29, an ensemble-based pipeline, to call structural variants (SVs) in 1,285 sequenced Samoans across all chromosomes.^3^ We excluded 9 samples: 3 due to insufficient quality to call SVs, 2 due to outlier autosomal copy numbers, and 4 due to outlier copy number variants (CNVs), resulting in high-quality SV calls for 1,276 participants (> 99% of WGS samples).

We called the following types of SVs: CNVs, complex SVs, deletions, duplications, insertions, inversions, and translocations. CNVs in this dataset all had multiallelic copy numbers. Complex SVs were either dispersed duplications or SVs with more than one SV type, such as a deletion and inversion. Structural variant calls were stored in Variant Call Format (VCF) files, where the reference allele was encoded as “N” and the alternate allele was encoded as the variant type, e.g., “<DUP>” for duplications and “<DEL>” for deletions. Similarly to SNVs, SV start positions were documented in the position column. Since all SVs were greater than 50 base pairs (bp), length and end positions were documented in the “INFO” column. With the exception of CNVs, genotypes were documented as 0/0, 0/1, or 1/1 for homozygous reference, heterozygous, and homozygous alternate genotypes, respectively, consistent with how SNVs are typically documented in VCF files. CNVs had alternative formatting because they were multiallelic; we marked the genotype field as missing and documented each participant’s copy number instead.

### Samoan-Specific SV and SNV (*Soifua Manuia-SV*) Reference Panel Generation

We created a Samoan-specific reference panel, henceforth known as the *Soifua Manuia-SV* reference panel, using the genotypes from 1,276 Samoans with WGS data and SV calls. The panel is composed of polymorphic variants on chromosomes 1 through 22 and X with one of the following quality filters: PASS, DUP2, MIS2, or TRI2 (Table S1). We set the minor allele count (MAC) threshold to 1 to retain rare variants, but we removed variants that were monomorphic in our sample to reduce computational time.

We did not explicitly exclude any SVs due to size, but we manually checked SVs greater than 20 Mb to ensure they appeared to be true signals and were biologically plausible. Additionally, all SVs retained in the panel had a no-call genotype rate of less than 5%. CNVs were dropped from the reference panel because their variable copy number format was incompatible with our imputation pipeline.

After filtering the data, we phased the reference panel using Eagle v2.4.1 and compressed it using Minimac v4.1.6.^26,27^ To handle male hemizygosity on chromosome X, we split chromosome X into four files: pseudo-autosomal region (PAR) 1, PAR2, non-PAR in females, and non-PAR in males.

### Imputation with the *Soifua Manuia-SV* Reference Panel

We applied this imputation pipeline to two groups of Samoans: Group A (*n* = 1,843) comprised Samoans who were genotyped with the Affymetrix 6.0 array and were not in the reference panel, whereas Group I comprised Samoans and American Samoans who were genotyped with the Illumina Global Screening array (*n* = 1,769) (Figure 1).

To align and harmonize the array data with the *Soifua Manuia-SV* reference panel, we used Genotype Harmonizer v1.4.28, dropping all array variants that were not in the reference panel.^28^ When converting from PLINK v1.9 to VCF format, we used BCFtools +fixref with hg38 fasta files from the UCSC Genome Browser to maintain allele formatting as reference/alternate rather than major/minor.^29–31^ Afterward, to phase the harmonized array data using the same reference panel, we used Eagle v2.4.1.^26^

Subsequently, to impute SNVs and SVs from the *Soifua Manuia-SV* reference panel into the Samoan array data, we used Minimac v4.1.6, designating “chunks” to span the entirety of each autosome to avoid splitting large SVs.^27^ Chromosome X was imputed in four distinct regions (PAR1, PAR2, non-PAR in females, and non-PAR in males), then the imputed variants were merged into a single file. We assessed imputation quality using Minimac’s r-squared (r^2^) metric, which estimates the squared correlation between the imputed genotypes and their unobserved, true genotypes. We considered imputed variants with an r^2^ < 0.3 to be poor quality, r^2^ ≥ 0.3 as having at least good quality, and r^2^ ≥ 0.8 as high quality.

### Imputation with a Samoan-Specific SNV-Only (Samoan-SNV) Reference Panel

Our original Samoan-specific reference panel, the *Soifua Manuia* panel, differs from the *Soifua Manuia-SV* panel in three ways: the original panel contained variants that were monomorphic in our sample, it did not have any structural variants, and it included nine more Samoans.^9^ To assess the impact of including SVs on SNV imputation performance, we generated an alternate version of the original panel, hereafter referred to as the Samoan-SNV panel, by removing the monomorphic variants and the nine additional participants to isolate the effects of SV inclusion. Then, we imputed SNVs across chromosomes 1 through 22 and X into Group A, using the imputation pipeline detailed above (Genotype Harmonizer v1.4.28, BCFtools, Eagle v2.4.1, and Minimac v4.1.6).^26–28^ Since Group A and Group I both only comprised Samoans, imputation performance was expected to be comparable; therefore, we only imputed Group A with the Samoan-SNV panel to reduce computational burden.

### Imputation with 1000 Genomes SV and SNV (1000G-SV) Reference Panel

As described above, Noyvert et al. created a publicly available, multi-ancestry reference panel comprising 888 participants from 1000G, containing both SVs and SNVs for chromosomes 1 through 22. They sequenced participants using Oxford nanopore long-read sequencing, aligned the data to hg38, and called SVs using Sniffles2 v2.0.7.^7^ We downloaded the full panel, henceforth referred to as the 1000G-SV panel, and used it to compare imputation performance between a multi-ancestry reference panel (the 1000G-SV panel) and a Samoan-specific reference panel (the *Soifua Manuia-SV* panel) for SNV and SV imputation in Samoans.

We did not make significant changes to this reference panel, as the genotypes were phased in our desired genome build and the variants were polymorphic. Noyvert et al. formatted most SVs in their VCF file as follows: the position column is the start position, the reference allele is “N,” and the alternate allele is the SV type and SV length, e.g., “<INS:97>” for a 97 bp insertion. Breakend variants differed from this format: since they are unresolved structural variants, where only one position is certain, their alternate allele lists the breakend identification number rather than an SV length. We split the VCF file by chromosome and compressed it into alternative file formats for faster phasing and imputation into our target datasets.

Using the same pipeline we created for the prior rounds of imputation, we again harmonized Group A genotypes with Genotype Harmonizer v1.4.28, fixed alleles with BCFtools, phased genotypes with Eagle v2.4.1, and imputed genotypes with Minimac v4.1.6, all with the 1000G-SV reference panel.^7,26–29^ As above, we did not use the 1000G-SV panel to impute genotypes into Group I as imputation performance was expected to be comparable across the two Samoan groups.

### Imputation Concordance

To assess SNV concordance, we compared imputed SNVs to sequenced-derived SNVs for nine Samoan participants using BCFtools gtcheck.^29^ These participants had WGS data, but they were excluded from the *Soifua Manuia-SV* reference panel due to insufficient quality to call SVs. We imputed these participants with each of the three reference panels as part of Group A. To calculate genome-wide SNV concordance, we divided the number of matching genotypes by the total number of matching sites, then we took the average across all nine participants. Of note, BCFtools gtcheck does not assess concordance at sites that are monomorphic in both the reference and query files. Additionally, since the Affymetrix 6.0 array omitted markers in PAR2 of chromosome X, we were unable to impute this region with any of the reference panels and thus could not compare the concordance of SNVs in this region.

We also assessed SNV and SV imputation concordance across arrays. During quality control, using empirical kinship matrices, we incidentally identified a participant who was recruited twice: once in American Samoa in 1990 and again in Samoa in 2010. This individual was genotyped on both arrays and was therefore imputed in Group A and in Group I using the *Soifua Manuia-SV* reference panel. Rather than immediately excluding the duplicate, we leveraged this as an opportunity to assess whether the choice of genotyping array scaffold influenced imputation results.

## Results

### Population Structure

To explore the population homogeneity of Samoans, we generated principal component (PC) and admixture plots comparing Samoans and American Samoans to population groups from 1000G (*n* = 2,504).^19,20^ The PC plots demonstrated that Samoans cluster together, regardless of polity, and that they represent a genetically unique group which is not adequately represented by 1000G (Figure S1).^19,20^ Notably, 95.2% of the genetic content across the Samoan participants is labeled as genetically dissimilar to the 1000G populations at *K* = 6 (Figure S2A). The admixture present is spread across the reference panel, Group A, and Group I participants (Figure S2B).

### Structural Variants

#### Samoan SVs Identified Using Short-Read WGS

In total we identified 47,644 unique SVs with high quality in the 1,276 Samoans with whole genome sequencing. Of these SVs, 18,595 were deletions, 17,972 were insertions, 10,034 were duplications, 564 were complex SVs, 414 were CNVs, 64 were inversions, and 1 was a reciprocal translocation. Overall, most SVs (78%) were between 50 bp and 500 bp long, and the longest SV was a 173 Mb inversion on chromosome 1. There were 11,111 variants that had a MAC of 1. Compared to SVs that were called in gnomAD v2.1 and lifted over to hg38, 24,612 (52%) of the SVs in Samoans were novel.^3,32^ On average we identified 7,986 unique SVs per participant.

#### SVs Incorporated into the Soifua Manuia-SV Reference Panel

Of the 47,644 SVs, we dropped all CNVs (*n* = 414) since their multiallelic formatting was incompatible with our imputation pipeline, all SVs on chromosome Y (*n* = 147), and all SVs that were homozygous in all participants (*n* = 18). We then reviewed three SVs that were over 20 Mb long: one translocation and two inversions. The translocation (MAC = 1) had breakpoints at chr9:130786065 (intron 1 of *ABL1* transcript NM_007313.3) and chr22:23290218 (intron 13 of *BCR*) and resembled the Philadelphia chromosome. We dropped this SV because the Philadelphia chromosome is typically somatic and is thus inappropriate for imputation.^33,34^ The two inversions were both pericentric with a MAC of 1 (173 Mb inversion on chromosome 1 and 38.4 Mb inversion on chromosome 7); we kept them in the reference panel because they appeared to be true signals that were biologically plausible. Read level investigation of each inversion revealed patterns consistent with a large inversion alone, without evidence supporting other structural variant types (e.g., insertions) or a more complex rearrangement. Large inversions are known to be compatible with life, but carriers may present clinically with infertility.^35,36^ In total, we incorporated 47,064 SVs into the *Soifua Manuia-SV* reference panel.

Figure 2 depicts the types, lengths, and frequencies of SVs in the *Soifua Manuia-SV* reference panel, where an allele frequency (AF) < 0.0005 corresponds to an allele count (AC) of 1. Of the SVs in this panel, 51% were unique to Samoans. Additionally, 1,118 SVs were rare in gnomAD (AF < 0.01) but were common in Samoans (AF > 0.05). In total, this reference panel had 23,069,811 variants, of which 47,064 were SVs (0.2%).

**Figure 2:**
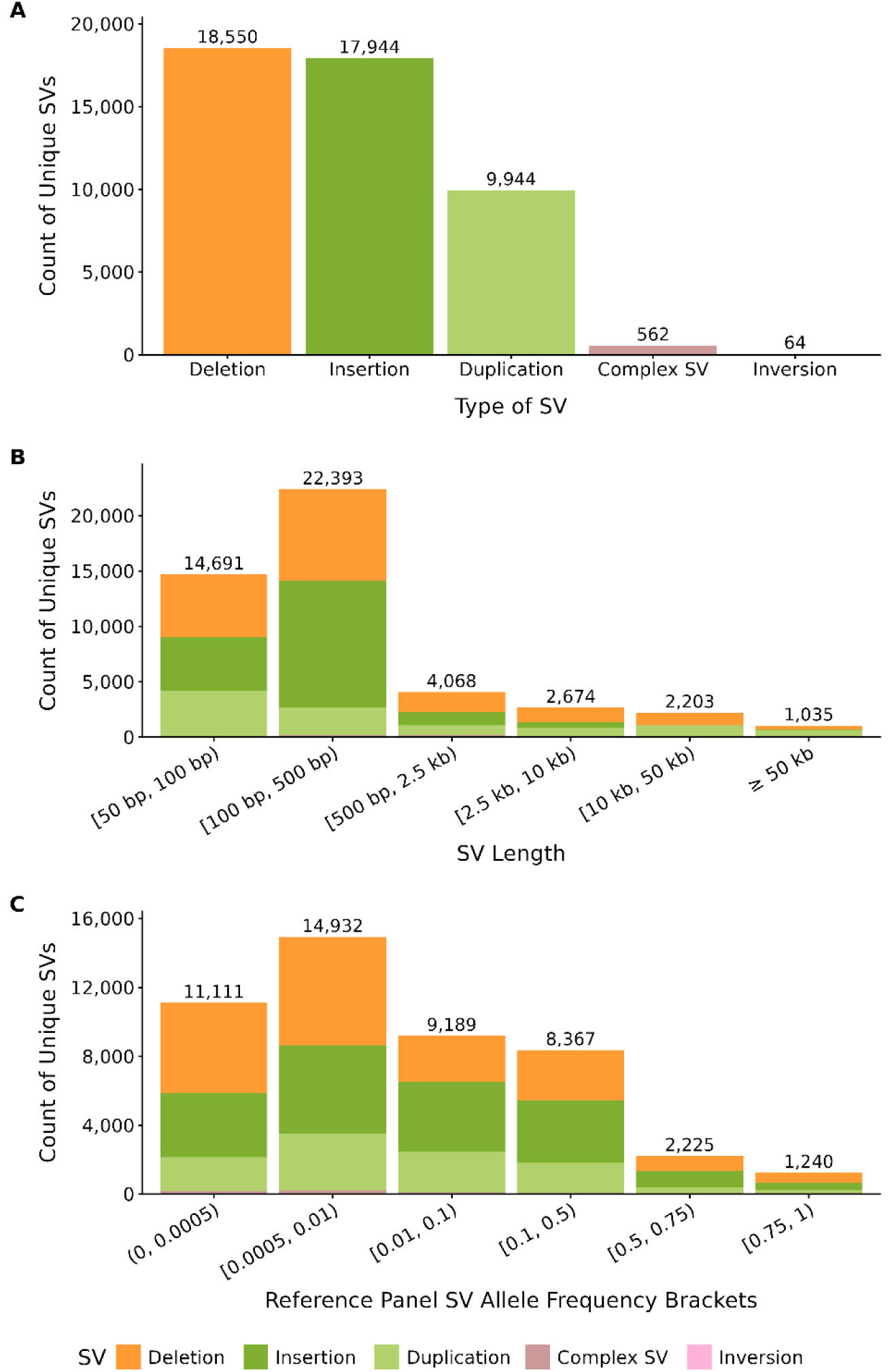
Structural variants in the *Soifua Manuia-SV* reference panel. A) SV types in the *Soifua Manuia-SV* panel. B) SVs in the *Soifua Manuia-SV* panel colored by SV type and split into bins by SV length. C) SVs in the *Soifua Manuia-SV* panel colored by SV type and split into bins by SV allele frequency.

### Reference Panels

Table 1 consolidates the similarities, dissimilarities, and variant counts between the *Soifua Manuia-SV*, Samoan-SNV, and 1000G-SV reference panels used for imputation. The 1000G-SV panel has 19.3 million more polymorphic variants than the *Soifua Manuia-SV* panel. In part, this is because the 1000G-SV panel used long-read sequencing to call SVs, which can typically detect more SVs than short-read sequencing. However, much of the difference is because the *Soifua Manuia-SV* panel comprised only Samoans, a founder population, whereas the 1000G-SV panel was multi-ancestry and was thus more genetically diverse. In total, we excluded 829 million SNVs from the Samoan-specific panels because they were monomorphic in our sample of 1,276 Samoans.

**Table 1:**
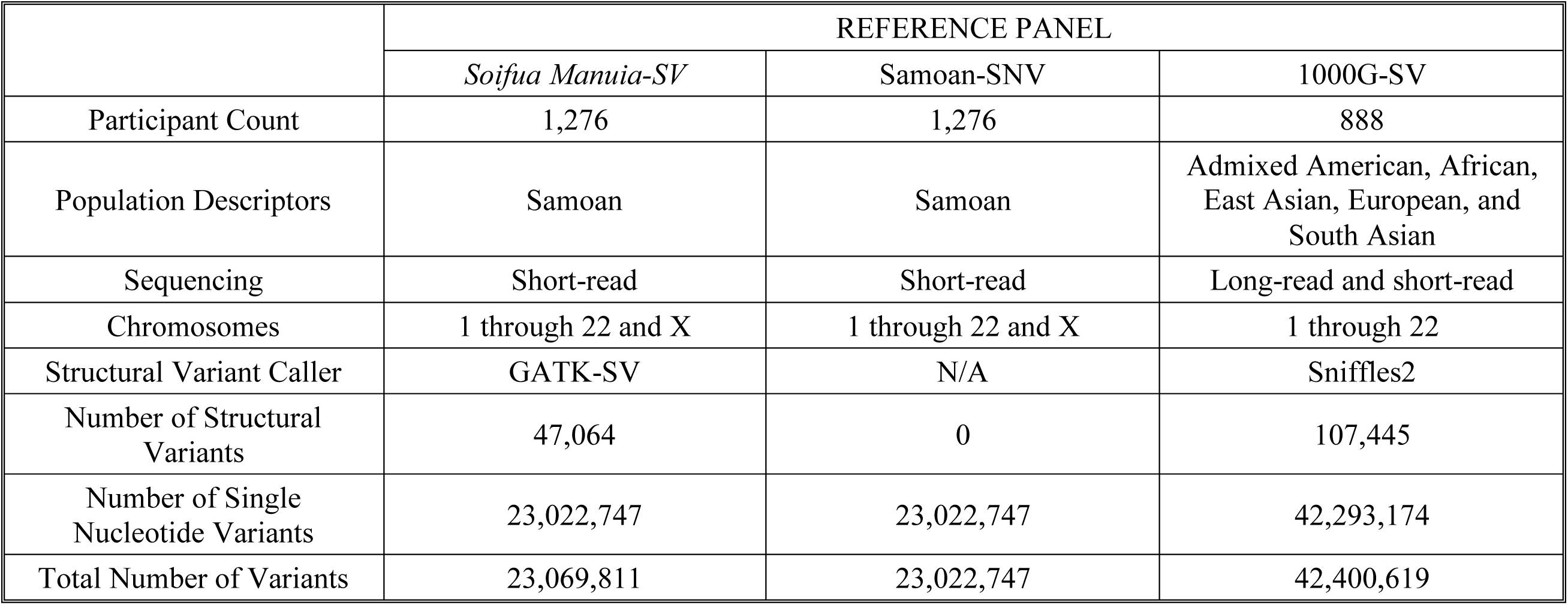
Reference panel overview. We excluded all variants that were monomorphic within each reference panel group, so each variant has a MAC ≥ 1.

|  | REFERENCE PANEL |  |  |
| --- | --- | --- | --- |
|  | *Soifua Manuia-SV* | Samoan-SNV | 1000G-SV |
| Participant Count | 1,276 | 1,276 | 888 |
| Population Descriptors | Samoan | Samoan | Admixed American, African, East Asian, European, and South Asian |
| Sequencing | Short-read | Short-read | Long-read and short-read |
| Chromosomes | 1 through 22 and X | 1 through 22 and X | 1 through 22 |
| Structural Variant Caller | GATK-SV | N/A | Sniffles2 |
| Number of Structural Variants | 47,064 | 0 | 107,445 |
| Number of Single Nucleotide Variants | 23,022,747 | 23,022,747 | 42,293,174 |
| Total Number of Variants | 23,069,811 | 23,022,747 | 42,400,619 |

### Imputation Concordance

Averaging across nine participants who were sequenced and imputed in Group A, SNVs imputed using the *Soifua Manuia-SV* reference panel were 99.63% concordant with sequenced SNVs across 23,019,996 SNVs on chromosomes 1 through 22 and X. Similarly, the Samoan-SNV panel yielded 99.66% concordance across the same 23,019,996 SNVs. We could not impute PAR2 of chromosome X since it was missing from the Affymetrix scaffold, so those SNVs were not evaluated in concordance metrics.

Of the 42,293,174 SNVs in the 1000G-SV reference panel, there was 98.87% concordance across 31,140,400 shared autosomal SNVs. The remaining 11 million SNVs in the 1000G-SV panel did not have an exact chromosome position and alternate allele match with the Samoan WGS SNVs, likely due to differences in sequencing methodology and quality control.

We also assessed SNV and SV concordance in the incidental cross-cohort duplicate who was imputed in both groups using the *Soifua Manuia-SV* panel. Across 23,067,048 shared variants, the variants imputed using the Illumina Global Screening array (Group I) were 99.62% concordant with the variants imputed using the Affymetrix 6.0 array (Group A), suggesting that the scaffolds perform similarly.

### Imputation Performance

#### R^2^ by Minor Allele Frequency

We visualized genome-wide mean and median r^2^ scores for all imputed variants to assess imputation accuracy across minor allele frequency bins. Figure 3 compares imputation between Group A and Group I when using the *Soifua Manuia-SV* panel, as well as compares how the *Soifua Manuia-SV* panel performs against the Samoan-SNV and 1000G-SV panels in Group A. We further split the plots to view trends for structural variants specifically (Figure 3).

**Figure 3:**
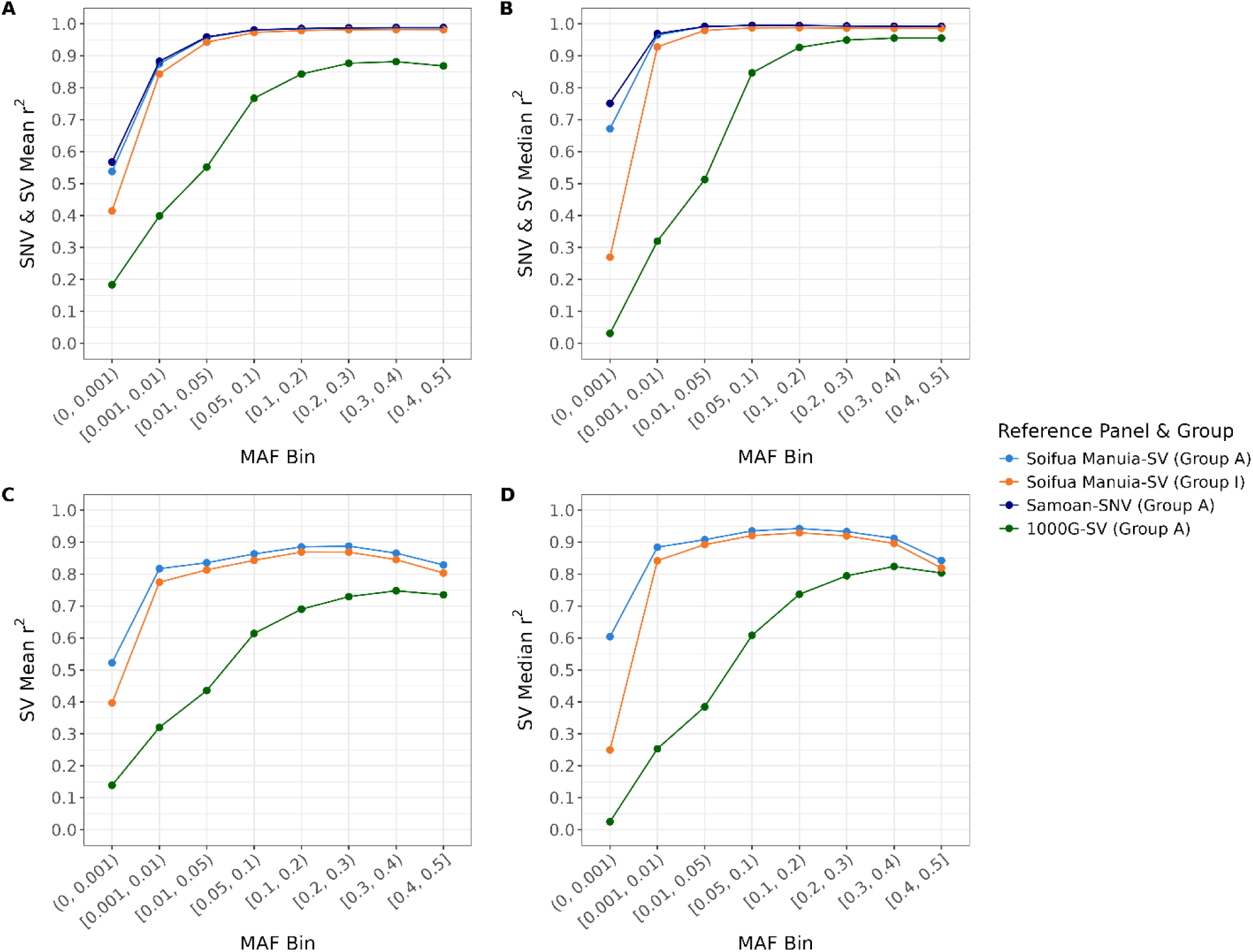
Genome-wide r^2^ by MAF bin. Plots A and B depict mean and median r^2^, respectively, for single nucleotide variants (SNVs) and structural variants (SVs) by minor allele frequency (MAF) bin for Group A, imputed using three different reference panels, and for Group I, imputed using the *Soifua Manuia-SV* panel. Plots C and D depict the mean and median r^2^, respectively, by MAF bin for SVs only; the Samoan-SNV panel is omitted from plots C and D because it does not contain any SVs. See Figure 1 for demographic and array information for each group.

#### Imputed Variants with High Quality Across Reference Panels

To compare imputation quality across reference panels and scaffolds, we imputed Group A using three different reference panels and Group I using the *Soifua Manuia-SV* reference panel. Table 2 highlights how these differences affect the number of high-quality imputed variants (r^2^ ≥ 0.8) and their average quality scores (r^2^). Group A had 9.5% more high-quality imputed variants than Group I, likely because its scaffold included 40.8% more genotyped SNVs. To visualize how effective each panel is at imputing rare variants, Figure 4 depicts the number of high-quality imputed variants in Group A by reference panel, stacked by MAF bin.

**Figure 4:**
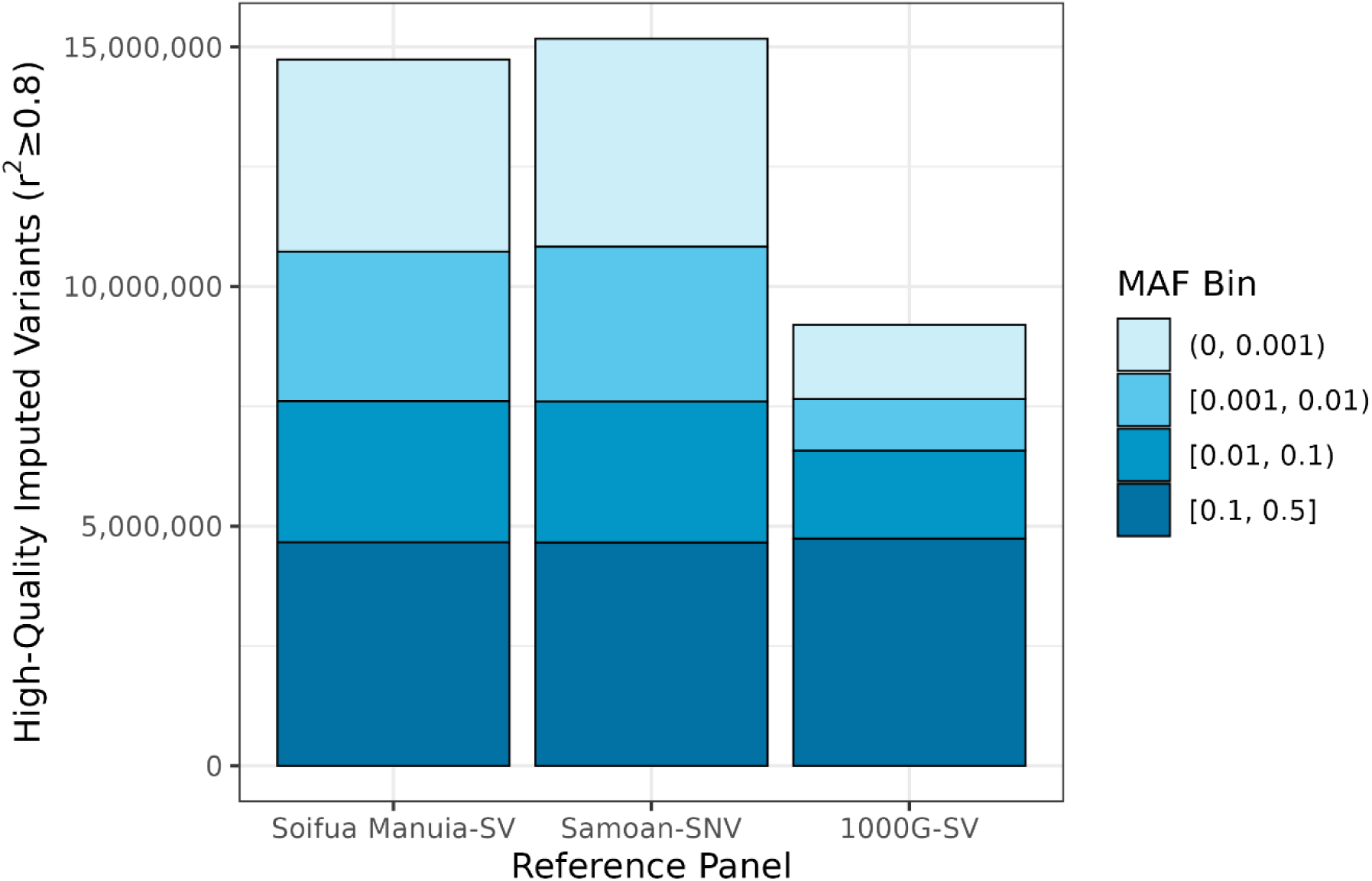
Count of high-quality imputed variants in Group A by reference panel and MAF bin. High-quality imputed variants (r^2^ ≥ 0.8) from each reference panel, colored by minor allele frequency (MAF) bin.

**Table 2:** High-quality imputation metrics. Imputation counts and statistics for high quality (r^2^ ≥ 0.8) imputed variants across two groups and three reference panels. Key: SNV = single nucleotide variant; SV = structural variant; Avg = average; N/A = not applicable.

| Reference Panel | *Soifua Manuia-SV* |  |  |  | Samoan-SNV |  | 1000G-SV |  |
| --- | --- | --- | --- | --- | --- | --- | --- | --- |
| Group | Group A |  | Group I |  | Group A |  | Group A |  |
| High Quality Variants (Count \| Avg $r^2$) | 14,735,723 | 0.97 | 13,456,959 | 0.96 | 15,170,397 | 0.97 | 9,200,036 | 0.94 |
| High Quality SNVs (Count \| Avg $r^2$) | 14,710,056 | 0.97 | 13,434,288 | 0.96 | 15,170,397 | 0.97 | 9,183,231 | 0.94 |
| High Quality SVs (Count \| Avg $r^2$) | 25,667 | 0.94 | 22,671 | 0.93 | N/A | N/A | 16,805 | 0.92 |

### Imputed Good- and High-Quality Structural Variants

After removing all monomorphic and all poorly imputed (r^2^ < 0.3) structural variants, 81% (*n* = 38,126) of the SVs in the *Soifua Manuia-SV* reference panel were imputed with at least good quality (r^2^ ≥ 0.3) in Group A, compared to 44% (*n* = 46,804) of SVs in the 1000G-SV panel (Table 3). Regardless of SV type, SVs were well-imputed with the *Soifua Manuia-SV* panel (mean r^2^ range: 0.81-0.90) and had satisfactory but poorer imputation with the 1000G-SV panel (mean r^2^ range: 0.52-0.68) (Table 3). In Group I, we imputed 36,843 SVs (78%) with at least good quality using the *Soifua Manuia-SV* panel (mean r^2^ = 0.81, median r^2^ = 0.88).

**Table 3:** Imputed structural variants (SVs) with good and high quality (r^2^ ≥ 0.3) in Group A. Imputed counts and statistics only describe variants with good quality (r^2^ ≥ 0.3) and with a minor allele count ≥ 1. Key: bp = base pair; kb = kilobases; AF = allele frequency.

| Reference Panel | *Soifua Manuia-SV* |  |  |  |  | 1000G-SV |  |  |  |  |
| --- | --- | --- | --- | --- | --- | --- | --- | --- | --- | --- |
|  | Reference | Imputed | Imputed % | Mean r<sup>2</sup> | Median r<sup>2</sup> | Reference | Imputed | Imputed % | Mean r<sup>2</sup> | Median r<sup>2</sup> |
| TOTAL | 47,064 | 38,126 | 81.01% | 0.85 | 0.91 | 107,445 | 46,804 | 43.56% | 0.67 | 0.67 |
| <u>SV Type<sup>a</sup></u> |  |  |  |  |  |  |  |  |  |  |
| Deletion | 18,550 | 14,605 | 78.73% | 0.86 | 0.93 | 38,459 | 20,443 | 53.16% | 0.68 | 0.70 |
| Insertion | 17,944 | 14,981 | 83.49% | 0.85 | 0.91 | 59,953 | 24,799 | 41.36% | 0.66 | 0.66 |
| Duplication | 9,944 | 8,070 | 81.15% | 0.81 | 0.86 | 608 | 149 | 24.51% | 0.52 | 0.48 |
| Complex SV | 562 | 424 | 75.44% | 0.90 | 0.96 | - | - | N/A | N/A | N/A |
| Inversion | 64 | 46 | 71.88% | 0.86 | 0.92 | 5,729 | 701 | 12.24% | 0.56 | 0.51 |
| Breakend | - | - | N/A | N/A | N/A | 2,696 | 712 | 26.41% | 0.52 | 0.48 |
| <u>Length Bins<sup>b</sup></u> |  |  |  |  |  |  |  |  |  |  |
| [50 bp, 100 bp) | 14,691 | 12,356 | 84.11% | 0.84 | 0.88 | 42,824 | 19,929 | 46.54% | 0.66 | 0.65 |
| [100 bp, 500 bp) | 22,393 | 18,069 | 80.69% | 0.85 | 0.92 | 34,446 | 17,026 | 49.43% | 0.70 | 0.74 |
| [500 bp, 2.5 kb) | 4,068 | 3,318 | 81.56% | 0.88 | 0.95 | 13,287 | 5,168 | 38.90% | 0.65 | 0.63 |
| [2.5 kb, 10 kb) | 2,674 | 2,119 | 79.24% | 0.88 | 0.96 | 11,453 | 3,348 | 29.23% | 0.62 | 0.60 |
| [10 kb, 50 kb) | 2,203 | 1,579 | 71.67% | 0.75 | 0.76 | 2,301 | 531 | 23.08% | 0.56 | 0.52 |
| ≥ 50 kb | 1,035 | 685 | 66.18% | 0.74 | 0.75 | 438 | 90 | 20.55% | 0.49 | 0.42 |
| <u>AF Bins<sup>c</sup></u> |  |  |  |  |  |  |  |  |  |  |
| (0, 0.0005) | 11,111 | 4,754 | 42.79% | 0.81 | 0.89 | - | 3,554 | N/A | 0.65 | 0.63 |
| [0.0005, 0.01) | 14,932 | 12,543 | 84.00% | 0.83 | 0.90 | 55,575 | 11,049 | N/A | 0.58 | 0.52 |
| [0.01, 0.1) | 9,189 | 9,096 | 98.99% | 0.86 | 0.93 | 25,082 | 10,835 | N/A | 0.63 | 0.59 |
| [0.1, 0.5) | 8,367 | 8,319 | 99.43% | 0.90 | 0.94 | 17,643 | 13,441 | N/A | 0.76 | 0.82 |
| [0.5, 0.75) | 2,225 | 2,217 | 99.64% | 0.82 | 0.82 | 4,945 | 4,572 | N/A | 0.75 | 0.80 |
| [0.75, 1) | 1,240 | 1,197 | 96.53% | 0.71 | 0.69 | 4,200 | 3,353 | N/A | 0.63 | 0.62 |

<sup>a</sup> The *Soifua Manuia-SV* panel did not have any breakend variants that passed quality filters, and the 1000G-SV panel did not include any variants labeled as “Complex SV.”
<sup>b</sup> Since breakends are typically unresolved SVs, they are missing length information and were thus omitted from the “Length” section of the table.
<sup>c</sup> Each imputed SV with good quality maintained the same AF bin in Group A as in the *Soifua Manuia-SV* reference panel. The 1000G-SV reference panel did not include any variants with an AF of (0, 0.0005) due to its sample size of 888. The imputed % column is missing from the 1000G-SV AF section because the AF bins differed between the 1000G-SV panel and the imputed data.

Using the *Soifua Manuia-SV* panel, we imputed an average of 7,366 unique SVs per participant in Group A and 7,352 SVs per participant in Group I, for an overall average of 7,359 SVs per imputed participant. Since we had one cross-cohort duplicate, we removed the individual from Group I before calculating the average number of SVs. When using the 1000G-SV panel, we imputed an average of 13,873 SVs per participant in Group A.

Half of the SVs imputed with good- to high-quality in Group A using the *Soifua Manuia-SV* panel were novel (*n* = 19,143). Of the SVs that were also documented in other populations, 46.7% of SVs had a higher imputed Samoan allele frequency than overall allele frequency in gnomAD. For example, when imputed into Group A, 1,130 SVs that were rare in gnomAD (AF < 0.01) had an imputed Samoan allele frequency greater than 5%.^3,32^

## Discussion

The goal of this study was to detect SVs in a sample of more than 1,000 Samoans using short-read whole genome sequencing data and to accurately impute them in a sample of 3,611 Samoans with array data. We further sought to assess the extent to which SV inclusion impacts SNV imputation quality and whether population representation impacts SV and SNV imputation.

We detected 7,986 unique SVs per participant for a total of 47,644 SVs. Of the SVs, 52% have not been observed before and are potentially unique to Samoans or Pacific Islanders.^3,32^ In terms of SVs per genome, this is consistent with what has been observed in other populations at similar read depths and using a similar SV-calling pipeline.^3^ Additionally, the number of novel SVs compared to those observed in gnomAD is not atypical for a newly examined population.^3,37^ Including these novel SVs in an imputation panel will enable opportunities to study SVs which could not have been imputed with less-representative panels like the 1000G-SV panel. Additionally, more than 1,000 SVs were common in Samoans but were rare in gnomAD participants. Such differences in population allele frequencies can be leveraged to discover the phenotypic effects of a broader range of SVs akin to what has been observed with SNVs.^38^ Ultimately, structural variants that are uniquely common or novel in Samoans hold untapped potential for genetic discovery.

Using the *Soifua Manuia-SV* panel, we observed high concordance between WGS and imputed genotypes in nine participants with WGS that were not included in the reference panels, and we also observed high concordance in imputed genotypes in a duplicate participant genotyped on two different arrays. In total, we imputed over 5.5 million more high-quality variants and over 8,000 more high-quality SVs, all with a higher average r^2^, using the *Soifua Manuia-SV* panel compared to the 1000G-SV panel. Additionally, the *Soifua Manuia-SV* panel outperformed the 1000G-SV panel across the entire MAF range in both mean and median quality scores, with the greatest outperformance at the lowest MAFs. Furthermore, it outperformed the 1000G-SV panel in the number of high-quality SVs and SNVs imputed, and the mean and median r^2^ of good-quality SVs were 27% and 36% higher, respectively.

Most likely, these improvements are due to better haplotype matching, which is fundamental for accurate imputation. None of the participants in the 1000G-SV panel are Polynesian; given the low genetic similarity between 1000G participants and Samoan participants, as illustrated by our ADMIXTURE analyses, there should be poorer haplotype matching. This would explain why Samoans were overwhelmingly better imputed when using the Samoan-specific *Soifua Manuia-SV* panel as opposed to the 1000G-SV panel.

There were 3% fewer high-quality SNVs imputed using the *Soifua Manuia-SV* panel compared to the Samoan-SNV panel, but three quarters of these unimputed variants were very rare. One possible explanation for this is that SV inclusion may have split haplotypes carrying these rare alleles, therefore reducing imputation confidence but better representing the true haplotype diversity at these loci. As such, the Samoan-SNV panel was likely overconfident at imputing these very rare SNVs compared to the *Soifua Manuia-SV* panel.

SV length impacted imputation quality across both panels, with small SVs averaging higher r^2^ scores than large SVs (≥ 10 kb). Our imputation pipeline relies on accurate linkage disequilibrium (LD) estimates for genotype harmonization, phasing, and imputation. Both reference panels documented SVs at the start position of the variant, regardless of the size or type of SV. Given that some SVs are large enough to span multiple haplotype blocks (≥ 1 Mb), it is possible that coding the variant’s position at the middle or end of the SV might influence its haplotype block placement and thus impact imputation results. This is less likely to be an issue for small SVs. Concordantly, a study using Atlantic salmon to assess SV imputation with GLIMPSE software also found that SVs in the longest length bins had poorer positive predictive values than SVs in short length bins.^39^ Additionally, small SVs may be better tolerated than large SVs, influencing their prevalence through natural selection. In the *Soifua Manuia-SV* panel, 34% of the SVs at least 10 kb in length had an AC of 1, so some of these variants may be *de novo* or are otherwise very rare. This skew toward lower frequencies in larger SVs could also result in the lower r^2^ scores observed here.

While including SVs with an AC of 1 in a reference panel runs the risk of including *de novo* variants, excluding them diminishes our ability to detect rare variants. We found that 43% of SVs with an AC of 1 (AF < 0.0005) in the *Soifua Manuia-SV* panel were imputed with at least good quality (mean r^2^ = 0.81), even though imputation pipelines typically have reduced performance for rare variants. Similarly to SVs that are extremely rare, SVs that are extremely common also have lower r^2^ scores because the reference allele is rare. We coded the frequency bins in Table 3 by AF instead of MAF to highlight that both panels had thousands of SVs that were more common than the reference allele. In particular, 39% of SVs with an AF ≥ 0.75 in the *Soifua Manuia-SV* panel were novel in Samoans, yet they were prevalent enough to greatly outnumber the reference allele.

Since long-read sequencing can be used to identify at least twice as many SVs as short-read sequencing, there are likely many additional Samoan-specific SVs extant in this cohort yet unobserved in this study.^5^ Thus, a future project should include long-read sequencing on the Samoans with WGS, calling SVs on the long-read WGS data, constructing a new reference panel with the updated SV data, and re-imputing SVs into the remaining Samoan participants.

In addition to imputing genotypes in Samoan research participants, this panel has the potential to impute variants into genetically similar individuals of other Polynesian ethnicities. There is evidence that Samoans, Tongans, and Te Ggana Tuuvalu–speaking Tuvaluans have high genetic similarity, and therefore the *Soifua Manuia-SV* panel could be highly useful for imputing genotypes in Tongans and some Tuvaluans.^40^ There is less genetic similarity between these “West Polynesian” peoples and the “East Polynesian” peoples of French Polynesia and the Cook Islands.^40^ However, this difference is likely due to lower haplotype diversity, consistent with settlement from West to East Polynesian, and the additional diversity on the *Soifua Manuia-SV* panel should not diminish its use for imputation into shared haplotypes.^41^ Therefore, compared to imputation panels lacking any Polynesian representation, the panel could still have much greater utility for East Polynesian ethnicities, including, potentially, Native Hawaiians and Māori. Many Polynesians also have recent ancestry from Europe, Asia, and Africa, and any panel used for imputation would need to include cosmopolitan representation to achieve the greatest utility for admixed individuals.

By generating an SV-inclusive Samoan-specific reference panel, we show that population-matched imputation substantially improves both SNV and SV imputation performance relative to a multi-ancestry panel lacking Polynesian representation, while maintaining robust SNV performance. Together, these findings highlight the importance of genetic similarity for accurate SV imputation and establish a practical framework for integrating SVs into large-scale genetic studies of underrepresented populations. By enabling large-scale study of structural variation in an underrepresented population, this work advances both methodological inclusivity and opportunities for genetic discovery. More broadly, our findings emphasize that accurate and equitable genomics requires intentional investment in population representation alongside continued technical innovation.

## Data Availability

Whole genome sequencing data used for creating the Soifua Manuia-SV reference panel and Samoan array data used for imputation of Group A are available on dbGaP (accession numbers: phs000972.v5.p1 and phs000914.v1.p1). The Samoan/American Samoan data used for Group I imputation have not been deposited in a public repository because participants did not provide consent for data sharing. Upon publication, the reference panel and a VCF file of SVs that were homozygous in all reference panel participants will become available in dbGaP (accession number: phs000972.v5.p1). Sample code for the Soifua Manuia-SV reference panel creation and Group A imputation are available on GitHub: https://github.com/Lspor/imputation.

## Supplemental Information

Supplemental Information includes two figures and one table.

## Data and Code Availability

Whole genome sequencing data used for creating the *Soifua Manuia-SV* reference panel and Samoan array data used for imputation of Group A are available on dbGaP (accession numbers: phs000972.v5.p1 and phs000914.v1.p1). The Samoan/American Samoan data used for Group I imputation have not been deposited in a public repository because participants did not provide consent for data sharing. Upon publication, the reference panel and a VCF file of SVs that were homozygous in all reference panel participants will become available in dbGaP (accession number: phs000972.v5.p1). Sample code for the *Soifua Manuia-SV* reference panel creation and Group A imputation are available on GitHub: https://github.com/Lspor/imputation.

## Acknowledgements

We would like to thank the Samoan participants of the study, local village authorities, and the many Samoan and other field workers over the years. We acknowledge the Samoan Ministry of Health, the American Samoan Department of Health, the Samoa Bureau of Statistics, and the Samoan Ministry of Women, Community and Social Development for their support of this research. We give particular thanks to two research assistants, Melania Selu and Vaimoana Lupematisila, who contributed to the 2010 recruitment and continue to assist us in our work in Samoa.

Molecular data for the Trans-Omics in Precision Medicine (TOPMed) program was supported by the National Heart, Lung, and Blood Institute (NHLBI). Genome Sequencing for NHLBI TOPMed: Samoan (phs000972.v5.p1) was performed at New York Genome Center Genomics (HHSN268201500016C) and Northwest Genomics Center (HHSN268201100037C). Core support including centralized genomic read mapping and genotype calling, along with variant quality metrics and filtering were provided by the TOPMed Informatics Research Center (3R01HL117626-02S1; contract HHSN268201800002I). Core support including phenotype harmonization, data management, sample-identity QC, and general program coordination were provided by the TOPMed Data Coordinating Center (R01HL120393; U01HL120393; contract HHSN268201800001I). We gratefully acknowledge the studies and participants who provided biological samples and data for TOPMed.

This work was funded by the National Institute of Health grants R01HL093093 (STM), R01HL133040 (RLM), R01AG009375 (STM), R01HL052611 (MI Kamboh), R01DK059642 (STM), and R01DK055406 (RD). Genotyping was performed in the Core Genotyping Laboratory at the University of Cincinnati, funded by National Institutes of Health grant P30ES006096 (S Pinney).

