## Supplemental Information for "Structural Variant Imputation in Samoans Using a Population-Specific Reference Panel"

[1] Department of Human Genetics, School of Public Health, University of Pittsburgh, Pittsburgh, PA, USA
[2] Center for Genomic Medicine, Massachusetts General Hospital, Boston, MA, USA
[3] Program in Medical and Population Genetics, Broad Institute of MIT and Harvard, Cambridge, MA, USA
[4] Department of Environmental and Public Health Sciences, College of Medicine, University of Cincinnati, Cincinnati, OH, USA
[5] Naseri & Associates Public Health Consultancy Firm & Family Health Clinic, Apia, Samoa
[6] Department of Epidemiology and Center for Global Public Health, Brown University School of Public Health, Providence, RI, USA
[7] Lutia i Puava ‘ae Mapu i Fagalele, Apia, Samoa
[8] Oceania University of Medicine, Apia, Samoa
[9] National University of Samoa, Apia, Samoa
[10] Department of Chronic Disease Epidemiology, Yale School of Public Health, New Haven, CT, USA
[11] Department of Public Health, LBJ Tropical Medical Center, Faga‘alu, AS, USA
[12] Division of Endocrinology and Metabolism, Department of Medicine, University of Pittsburgh, Pittsburgh, PA, USA
[13] Department of Anthropology, Brown University, Providence, RI, USA
[14] Department of Biostatistics and Health Data Sciences, School of Public Health, University of Pittsburgh, Pittsburgh, PA, USA
[15] These authors contributed equally to this work.

**Figure S1: Principal component (PC) plots.** Principal component plots are multi-dimensional: PC1 is the x-axis, PC2 is the y-axis, and PC3 is the z-axis. Population Key: AFR = African; AMR = Admixed American; ASM = American Samoan; EAS = East Asian; EUR = European; SAS = South Asian; SM = Samoan. Note: A single individual was recorded as having both EUR and AFR ancestries (EUR,AFR).

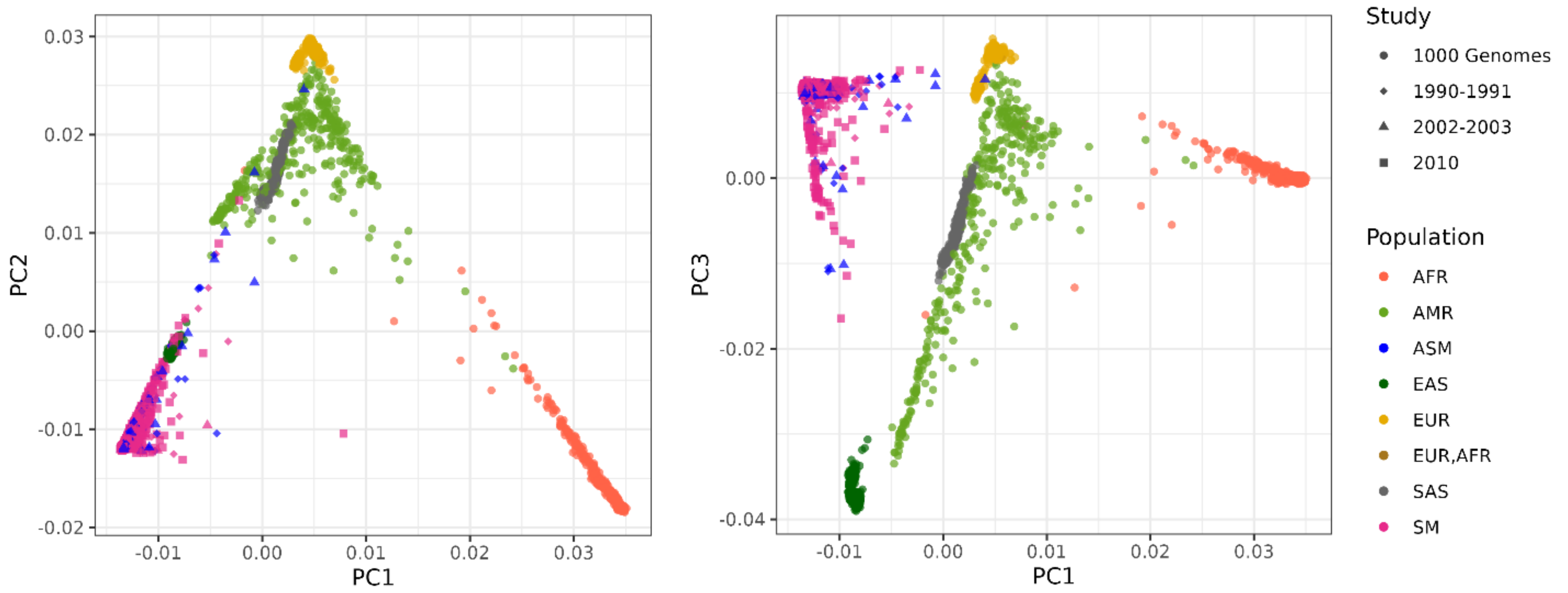

**Figure S2: ADMIXTURE plots.** Plot A depicts population structure for American Samoans (ASM), Samoans (SM), and 1000 Genomes populations —African (AFR), Admixed American (AMR), East Asian (EAS), European (EUR), and South Asian (SAS) — inferred using ADMIXTURE at $K = 6$. Plot B depicts population structure for the American Samoans and Samoans included in this study, split by the reference panel group, Group A, and Group I, with colors from Plot A.

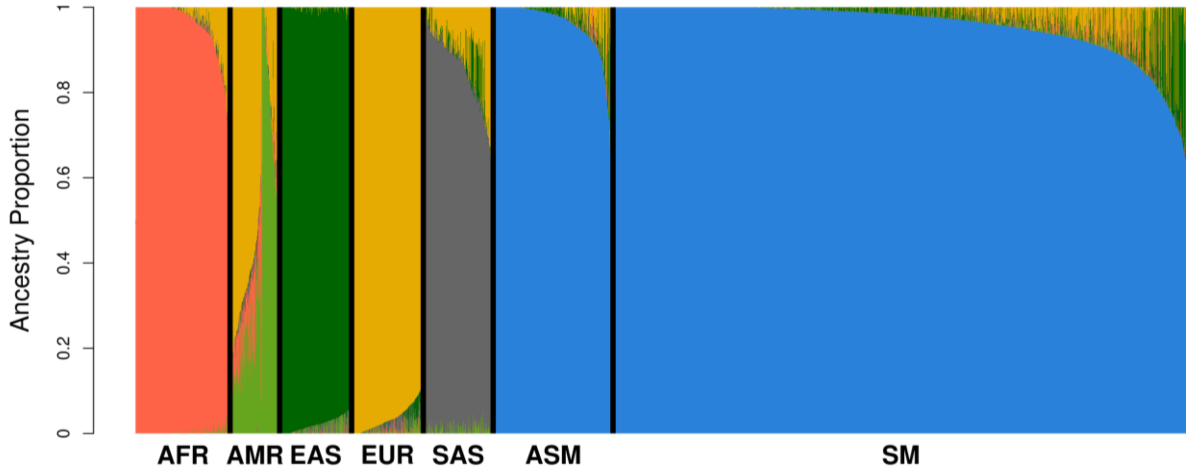


A)

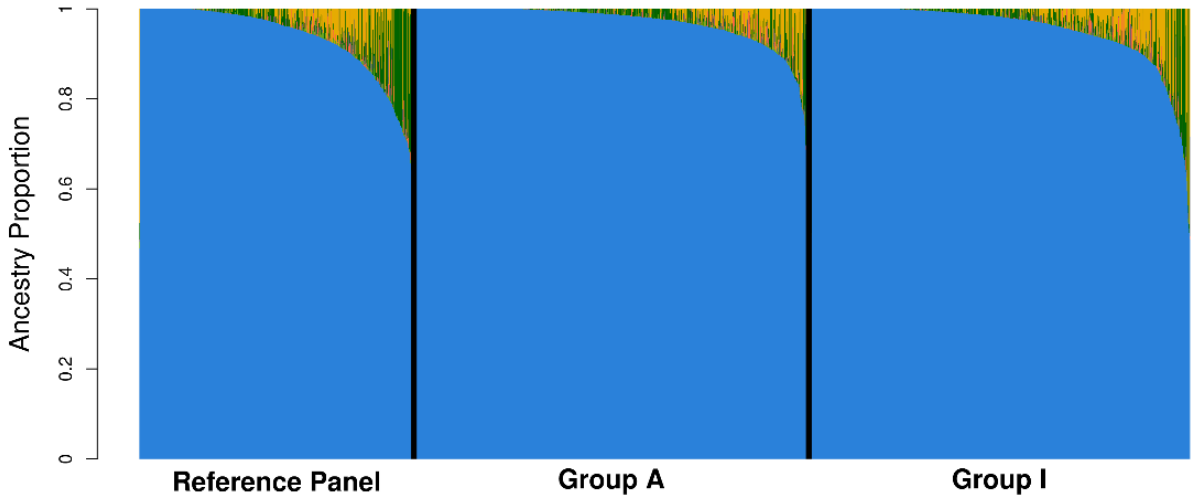


B)

**Table S1: VCF filter descriptions**

| Quality Filter | Quality Filter Description |
|---|---|
| PASS | All filters passed |
| DUP2 | Duplicate genotype discordance is greater than 2%, with at least two discordances |
| MIS2 | Genotype missing rate at depth 10 is greater than 2% |
| TRI2 | Mendelian genotype discordance is greater than 2%, with at least two discordances |
